# Identification of pronounced gender and geographic differences in the Diagnostic and Statistical Manual of Mental Disorders, fifth edition, text revision (*DSM-5-TR*)

**DOI:** 10.1101/2025.06.23.25330160

**Authors:** Alexa T. Diianni, Lauren C. Davis, Brian J. Piper

## Abstract

**Background:** Female representation in academia has increased in recent years, yet gender disparities among high-ranking positions remain prevalent. In academic psychiatry departments, the percentage of women holding high-ranking positions declines with increasing seniority, and previous research on the *Diagnostic and Statistical Manual of Mental Disorders*, fifth edition, revealed an underrepresentation of female authors. We have previously reported on financial conflicts of interest in the *Diagnostic and Statistical Manual of Mental Disorders*, fifth edition, text revision (*DSM-5-TR*), concluding that nearly 60% of authors received payments from industry, totaling to >$14.2m. Although the *DSM-5-TR* has international representation, geographic representation within the United States (US) has not been reported. In this study, we evaluated the gender and geographic location (US) of the *DSM-5-TR* task force, panel, and cross-cutting review group members, with respect to financial conflicts of interest.

**Methods:** We manually entered the names of *DSM-5-TR* task force, panel, and cross-cutting review group members in the National Provider Identifier (NPI) registry to obtain gender and geographic information, and in Open Payments to determine the type and amount of compensation received from 2016-19.

**Results:** There were 116 contributors (five task force, 83 panel, 28 cross-cutting review group members) that met the inclusion criteria. The 65 contributors (56.5%) received $14.6m from industry. Males accounted for >70% of contributors receiving funding and received >90% of total compensation. Three states were represented by over three-quarters (76.1%) of contributors.

**Conclusion:** Females accounted for less than one-third of all *DSM-5-TR* contributors and had fewer financial ties to industry. One of the five task force contributors was female. Over three-quarters of contributors represented three states while twenty-three states were not represented. To promote equitable gender and geographic representation in the *DSM-6*, the equal inclusion of male and female contributors, and contributors from various regions of the US, should be prioritized.

**Strengths and Limitations:**

- A primary strength of this study is that it considers the gender and geographic information of *DSM-5-TR* contributors in relation to financial conflicts of interest, which has not previously been reported.
- Our investigation sheds light on the disproportionate gender and geographic representation of *DSM-5-TR* authors.
- This study used the Open Payments database and NPI registry, which consist of mandated and self-reported disclosures, and it is thus possible that information obtained from these sources may have been outdated or absent.
- Only US states were considered for geographic analysis despite multiple contributors residing outside the US.
- Although there was a subset of contributors who hold medical licensure in multiple US states and/or have medical practices residing in more than one state, only the state of contributors’ primary medical practice was considered in the geographic analysis.

## Background

Female representation in academia has increased in recent years, yet gender disparities among high-ranking positions remain prevalent. The Association of American Medical Colleges reports that nearly half of all medical education faculty are female; however, women account for less than 30% of full professors and only one-fourth of department chairs (1). In academic psychiatry departments, female representation in senior positions exceeds the national average, though the percentage of women holding high-ranking positions declines with increasing seniority (1,2,3). Indeed, women have been historically underrepresented in clinical psychiatry, with female psychiatrists accounting for only 41.8% of all psychiatrists in 2023 in the United States (US) (4).

The *Diagnostic and Statistical Manual of Mental Disorders (DSM)*, published by the American Psychiatric Association, establishes a standard for symptom criteria and classifies psychiatric disorders (5). Often referred to as the “bible” of psychiatry, the *DSM* is used by health professionals and researchers in the US and abroad (6), and it is largely influential in the approval of new psychiatric medications (5). Previous research on the *Diagnostic and Statistical Manual of Mental Disorders*, fifth edition, revealed a disproportionate underrepresentation of female authors (7).

Earlier editions of the *DSM* have been criticized for insufficient transparency of financial ties to industry (5,8,9). Although financial conflicts of interest are not evidentiary of any wrongdoing, there is a wealth of literature demonstrating that they can create implicit bias and promote pro-industry habits of thought (10,11,12). As such, the National Institutes of Medicine (US) recommends that guideline development groups should be free of commercial ties (13). We have previously reported on financial conflicts of interest among the *Diagnostic and Statistical Manual of Mental Disorders*, fifth edition, text revision (*DSM-5-TR*) and found that nearly 60% of authors received payments from industry, totaling to >$14.2m (5).

It is widely acknowledged that notable differences exist among various geographic regions in the US, such as lifestyle factors, social and environmental determinants of health, and healthcare accessibility (14,15). Additionally, previous research indicates that the geographic location of upbringing has profound impacts on personality traits, emotional states, and community values, and it also acts as a predictor for mental health issues linked with social and environmental influences (16). Although the *DSM-5-TR* is represented by internationally based contributors, geographic representation within the US has not been reported (17). In this study, we evaluated the gender and geographic location (US) of the *DSM-5-TR* task force, panel, and cross-cutting review group members, with respect to financial conflicts of interest.

## Methods

### Procedures

In our previous study, the inclusion criteria consisted of *DSM-5-*TR panel and task force members who were based in the US and held physician (MD or DO) credentials (5). For the present study, the inclusion criteria was expanded to include contributors of the four *DSM-5-TR* cross-cutting review groups (i.e., culture, sex and gender, suicide, forensics) and the ethnoracial equity and inclusion review group (5,18). The cross-cutting review groups evaluated material pertaining to their specific expertise within each chapter, and the ethnoracial equity and inclusion review group examined all texts to ensure the usage of nondiscriminatory and non-stigmatizing language (19). For simplicity, the ethnoracial equity and inclusion review group was combined with the four cross-cutting review groups in this analysis. The addition of these groups was implemented to encompass a more extensive group of contributors, thereby providing a more inclusive depiction of representation in the *DSM-5-TR*.

To determine gender and geographic location (state of primary medical practice) of the *DSM-5-TR* task force, panel and cross-cutting review group contributors, each contributor’s name was manually entered in the National Provider Identifier (NPI) registry (20).

The *Physician Payment Sunshine Act* was passed in 2013, mandating all pharmaceutical and device manufacturers to report payments made to physicians and teaching hospitals (21). Subsequently, The Centers for Medicare and Medicaid Services developed Open Payments, a publicly accessible database that quantifies and categorizes payments given by industry to individual physicians and teaching hospitals (21). The name of each cross-cutting review group contributor was manually entered into Open Payments to determine the type and extent of compensation received during the years 2016-19. This period was selected to include the year in which development of the *DSM-5-TR* began (19) and the three preceding years, a period consistent with our previous study and the American Psychiatric Association’s disclosure policy for the *DSM-5* (5,9,22). Middle initials, NPI numbers, and location (state) were cross-referenced with the NPI registry to confirm the correct individual was identified in both systems.

Open Payments categorizes physicians’ compensation into ten types of payment: research payments, associated research funding, food and beverage, travel and lodging, consulting fee, other compensation, honorarium, education, gift, and ownership and investment (23). For the present study, the research payments category was combined to include both associated research funding and research payments.

Individuals without entry in Open Payments were coded as receiving no compensation, and individuals without entry in the NPI registry were excluded from this analysis. Since Open Payments and the NPI registry are dynamic databases, May 14, 2024, and April 30, 2024, were selected as cutoff dates for no further consideration of data, respectively. The Geisinger IRB deemed this not human subjects’ research, and the STROBE checklist for cross-sectional analyses was used in the creation of this report.

### Data-analysis

Descriptive analysis and formulation of heat maps were completed with Microsoft Excel. Other figures were constructed with GraphPad Prism version 10.4.2. A p < .05 was considered statistically significant.

## Results

After duplicate names were removed, 225 physicians were identified as contributors to the *DSM-5-TR* task force, panels, and cross-cutting review groups. There were 116 contributors (five task force, 83 panel, 28 cross-cutting review group members) that met the inclusion criteria of being a US-based physician with an active NPI and industry payments tracked in the Open Payments database (fig 1). Over two-thirds of the contributors were male (69.8%), and the rest were female (30.2%).

**Figure 1:**
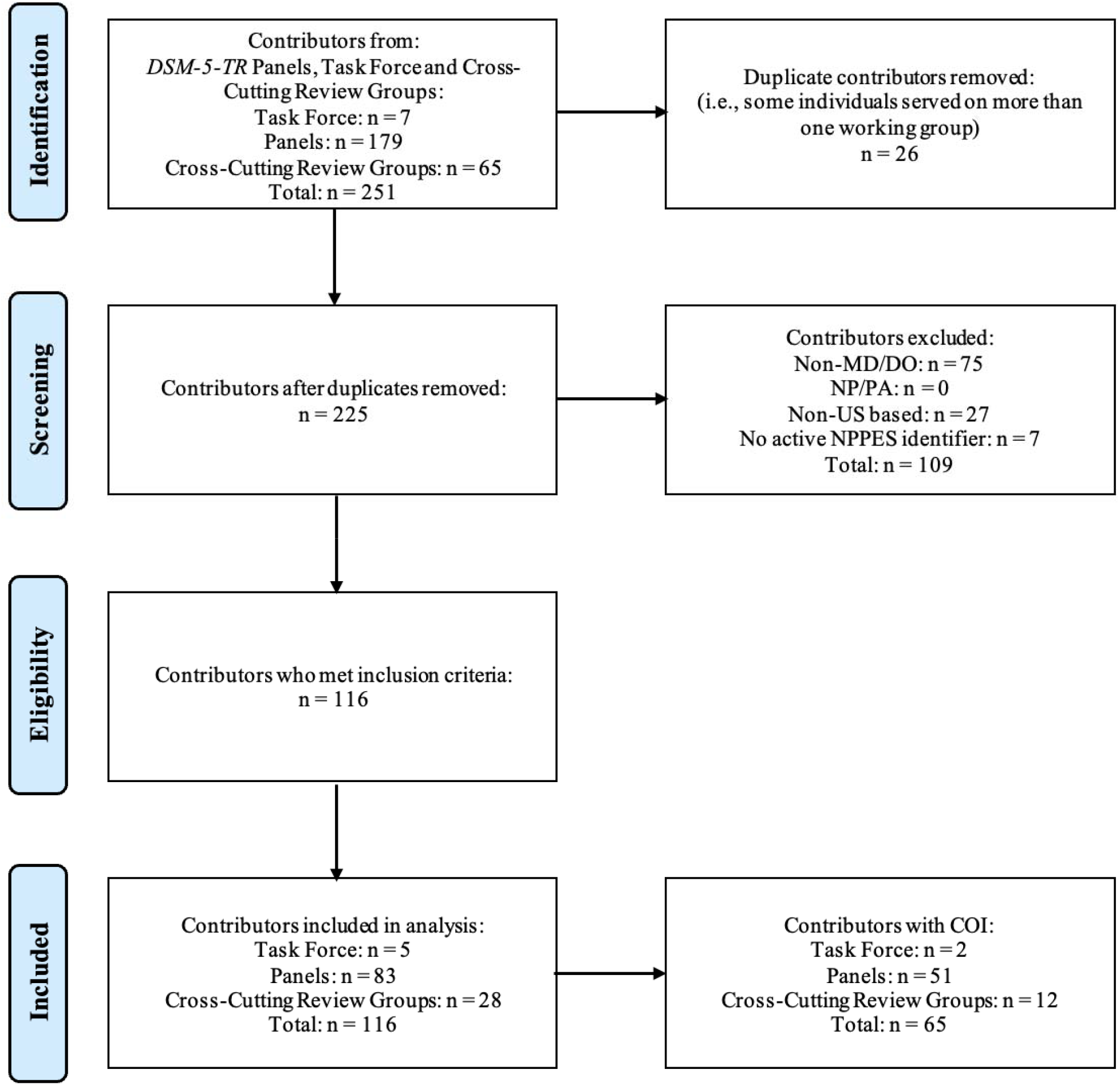
Flowchart of physician contributors identified from *Diagnostic and Statistical Manual of Mental Disorders*, fifth edition, text revision (DSM-5-TR) panel, task force and cross-cutting review groups members. Contributors were screened based on physician status (Doctor of Medicine or Doctor of Osteopathic Medicine) and being a resident of the US. Contributors who could not be identified in the National Provider Identifier (NPI) registry were also excluded. Four contributors served both on a panel(s) and cross-cutting review group(s) and were classified as panel members since panels are given higher authority than cross-cutting review groups in the revision process (18).

Contributors’ primary medical practices were unevenly distributed across the US. Just three states: New York (20), California (17), Massachusetts (14), accounted for three-quarters of contributors (51, 76.1%), and twenty-three states had zero contributors (fig 2). Sixteen states were represented by at least one female contributor.

**Figure 2:**
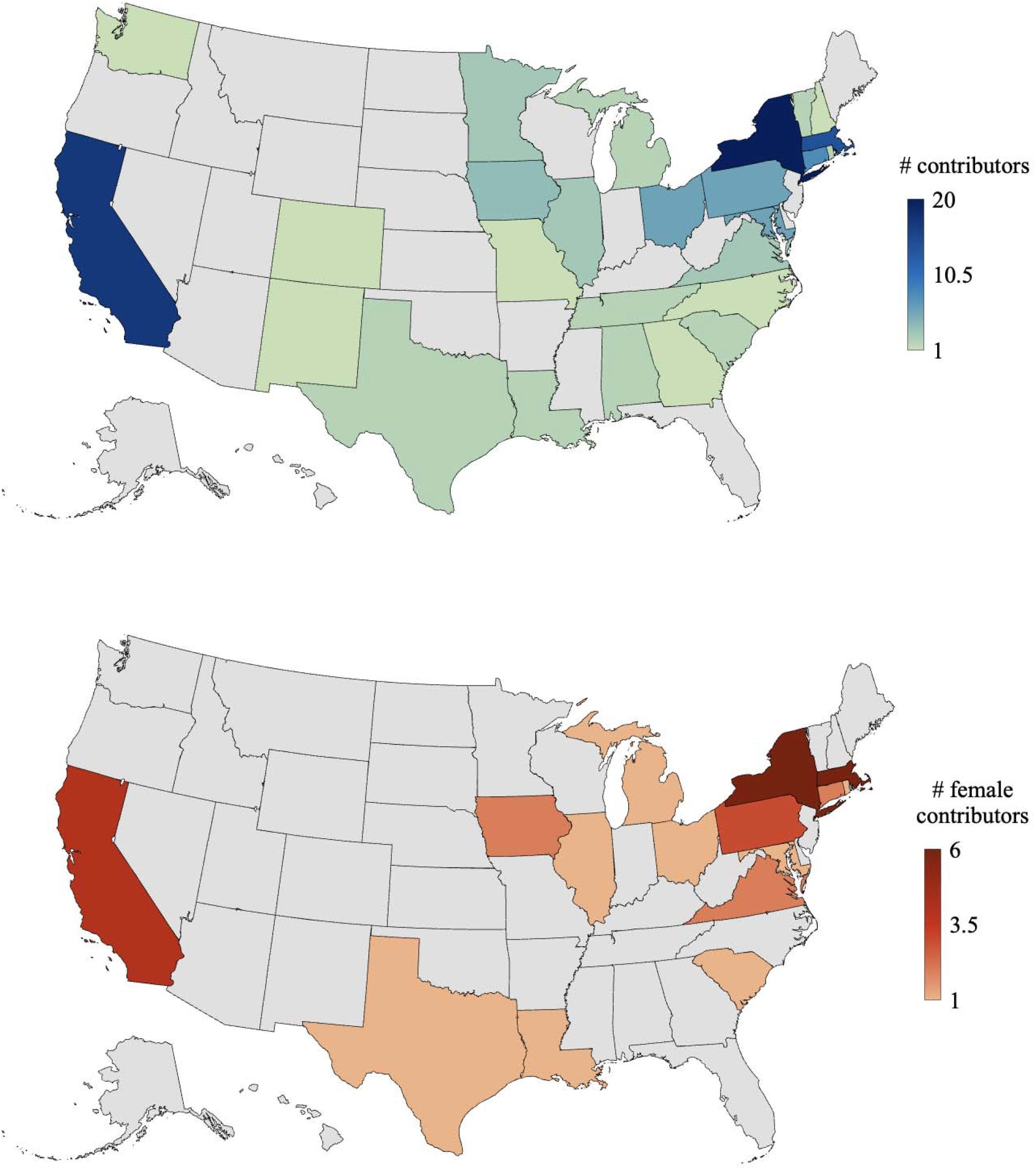
Non-homogenous distribution of the location of contributors to the Diagnostic and Statistical Manual of Mental Disorders, fifth edition, text revision (DSM-5-TR). The top map shows the locations of all contributors to the DSM-5-TR. States with the largest number of contributors are shown in dark blue, and states with the fewest number of contributors are shown in green. The bottom map shows the locations of female contributors to the DSM-5-TR. States with the largest number of female contributors are shown in dark red, and states with the fewest number of female contributors are shown in orange. In both maps, states without representation of contributors are shown in gray.

Nearly three-fourths (73.1%) of task force, panels, cross-cutting review groups included at least one female contributor (fig 3). Female contributors accounted for at least half of group membership in 7 of 26 groups (26.9 %) (i.e., sex and gender review (12/12), anxiety disorders (1/1); sexual disorders (2/3), obsessive-compulsive disorders (3/5), ethnoracial equity and inclusion (5/10), disruptive, impulse-control and conduct disorders (1/2), dissociative disorders (2/4)).

**Figure 3:**
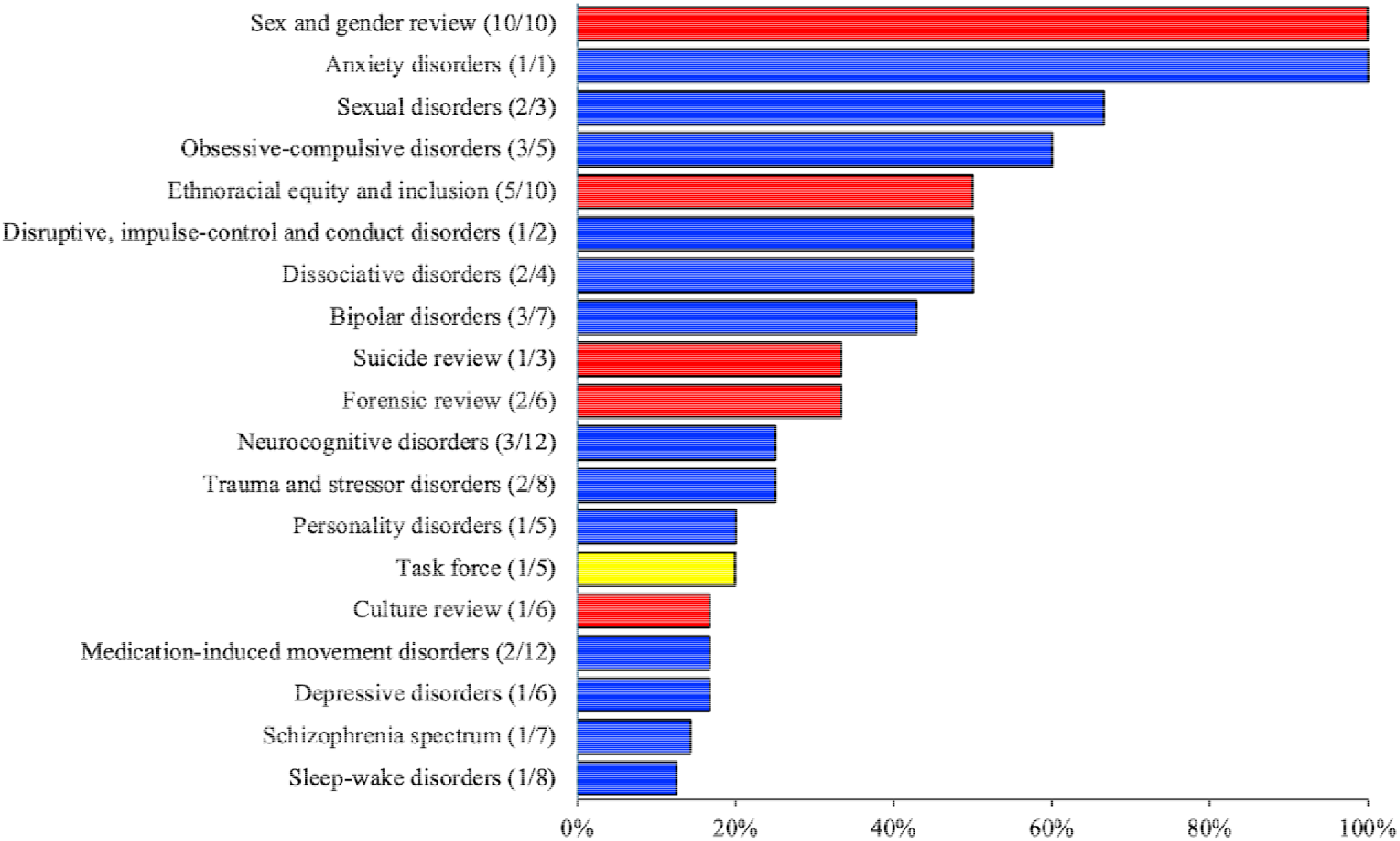
Proportion of female authors contributing to the task force, panels and cross-cutti review groups for the *Diagnostic and Statistical Manual of Mental Disorders* fifth edition, text revision *(DSM-5-TR)*. The proportion of female contributors is indicated in parentheses beside each group. Cross-cutting review groups are indicated in red, panels are indicated in blue, and task force is indicated in yellow. The seven panels for which no female authors contributed (i.e., neurodevelopmental disorders, somatic symptom disorders, feeding and eating disorders, elimination disorders, gender dysphoria, substance-related and addictive disorders, and paraphilic disorders) are not shown.

Sixty-five contributors (56.0%) had financial ties to industry, totaling to $14.6m. The 46 male contributors (70.8%) received a total of $13.2m, which represents over 90% of all payments made to *DSM-5-TR* contributors. On average, male contributors received a total of $287k, while female contributors received $72k. Thus, males received payments from commercial entities nearly four-times greater than those received by females.

The most common of the ten types of payments as listed in Open Payments were food and beverage (93.5% male, 51.4% female; total $95,948.61), travel and lodging (67.4% male, 28.6% female; total $699,782.77), and consulting (65.2% male, 28.6% female; total $1,213,835.83). The greatest difference in the proportion of male and female contributors receiving remuneration was in the honorarium category (6 male:1 female), followed by education (4.3 male:1 female), and research payments (3.4 male:1 female) (fig 4).

**Figure 4:**
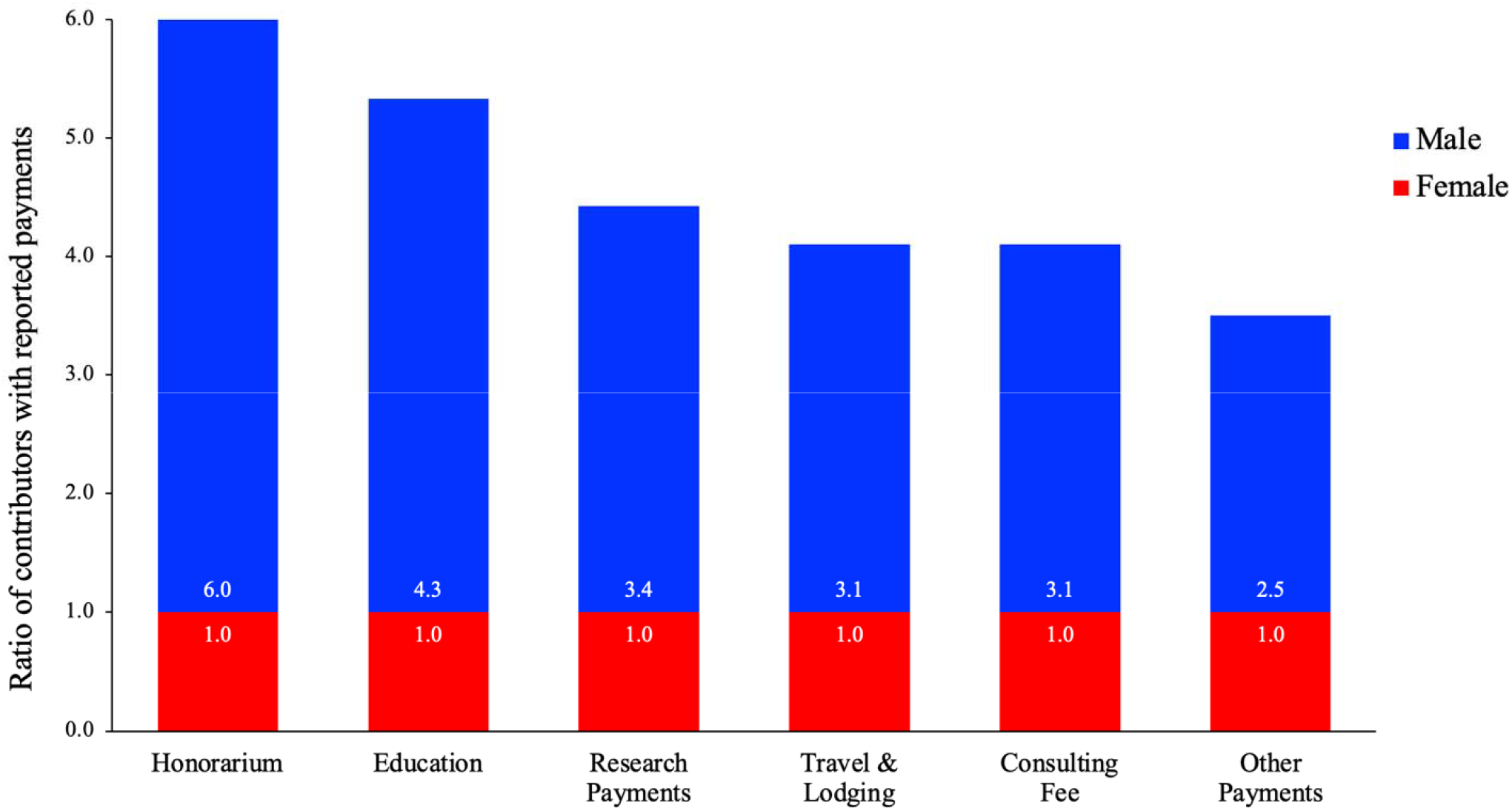
Ratio of male and female task force, panel and cross-cutting review group members of the American Psychiatric Association’s *Diagnostic and Statistical Manual of Mental Disorders*, fifth edition, text revision (*DSM-5-TR)* with reported compensation in each category of the Open Payments database from 2016-19. The research payments category includes both associated research funding and research payments, as provided in the Centers for Medicare and Medicaid Services’ Open Payments database.

## Discussion

In this study, we found that nearly 70% of contributors to the *DSM-5-TR* were male. Although 30% of female contribution is a step in the right direction, there is still work to be done considering approximately 40% of psychiatrists were female in 2023 (4). The decrease in female representation among high-ranking positions in medical academia has been termed the “leaky pipeline” (1,2,3). Consistent with this analogy, only one of the five (20%) task force contributors was female. It has been postulated that a lack of mentorship and networking opportunities for women may contribute to the gender disparities within senior roles in medical academia (2). Therefore, it is necessary to promote equal representation of male and female contributors within the task force of future *DSM* editions.

Moreover, previous research has shown that female patients seek psychiatric care and report receiving treatment at higher rates than male patients (24,25). It was reported that 63.2% of females reported discussing mental health concerns with their primary care provider compared to 54.2% of males (24). A report by the US Centers for Disease Control concluded that 28.6% of women received mental health treatment in 2021 compared to 17.8% of men (25). The increased proportions of female psychiatry patients necessitate equitable representation among *DSM* contributors. Accurately representing the patients that the *DSM* serves to diagnose enhances communication and trust between patients and their mental health specialists (7).

There were seven panels in which none of its contributors were female (i.e., gender dysphoria, neurodevelopmental disorders, somatic symptoms disorders, feeding and eating disorders, elimination disorders, substance-related and addictive disorders, and paraphilic disorders). The lack of female representation among these specific panels is notable, considering that somatic symptom disorders and feeding and eating disorders affect female patients at higher prevalence than male patients and would therefore benefit from the input of female contributors (7,18). This finding is congruent with an earlier report that found that among the authorship of two textbooks widely used in training physicians, *Katzung’s Basic and Clinical Pharmacology* and *Harrison’s Principles of Internal Medicine*, where males outnumbered females by about four-fold (26).

This investigation also found that female authors were both less likely to have conflicts of interest and, when they did, they were of smaller magnitude. There were 49 male contributors who received a total of $13.2m in industry payments, representing >90% of all financial conflicts of interest in the *DSM-5-TR*. The average of payments received from 2016-19 by male contributors was nearly four-times greater than that of female contributors. The reduction in conflicts of interest among females is certainly positive and is also observed within other medical specialties. Pronounced gender disparities in industry relationships were observed previously for radiation oncologists where males accounted for 72 and females accounted for none of the royalty recipients (27). Similarly, the top ten most highly renumerated contributors to UpToDate received $56.1 million and all ten were males (28).

There were fifteen male contributors and six female contributors who received “compensation for services other than consulting, including serving as faculty or as a speaker at a venue other than a continuing education program”. Male contributors received >95% of all funding in this category, totaling to $1.7m (supplemental fig 1). The “other compensation” category encompasses what is known as a key opinion leader amongst the pharmaceutical industry - “physicians who influence their peers’ medical practice, including but not limited to prescribing behavior” (29). The role of a key opinion leader is rooted in marketing and is widely acknowledged as a conflict of interest (5,30). Individuals who have participated on pharmaceutical companies’ Speakers Bureaus and thus have effectively worked as a key opinion leader, should be prohibited from *DSM* panel membership. Additionally, we recommend that the APA reinstates its disclosure policy for future editions of the *DSM*.

Furthermore, it is essential to ensure the perspectives expressed in a diagnostic manual are representative of the population intended for diagnosis (7). While the *DSM-*5-*TR* includes international representation (17), it is also important to consider the geographic representation of contributors within the US, given the diverse environmental, societal and psychological factors that impact mental health across the country (14,31,32). Three states, New York, California, and Massachusetts, were overrepresented and accounted for three-quarters of the *DSM-5-TR* authors that met inclusion criteria despite only accounting for one-fifth (20.0%) in 2019 of the US population (33). It is also noteworthy that the *DSM-5-TR* had no contributors from some of the more racially and ethnically diverse states (e.g. Hawaii and Florida) or from many rural states (e.g. Alaska and Oklahoma) (34).

### Strengths and Limitations

A primary strength of this study is that it considers gender and geographic information of *DSM-5-TR* authors in relation to conflicts of interest, which has not previously been reported. Our investigation serves to shed light on the disproportionate representation of *DSM-5-TR* authors. We hope this work will increase transparency and promote equity in authorship of future *DSM* editions.

Similar to our earlier report, the present study used information provided in the Open Payments database and NPI registry, which does not include payment and demographic information for physicians outside the US (5). Of the 225 contributors, 116 met the inclusion criteria, and thus it is possible that the 109 excluded contributors were free of industry ties. Furthermore, the National Provider Identifier registry is a self-reporting database in which physicians are responsible for updating their own NPPES data (20). Therefore, it is possible that a subset of information provided in the NPI registry is outdated or absent.

Although the *DSM-5-TR* has international representation among its contributors (18), only US states were considered for geographic analysis in this report due to the aforementioned limitations of Open Payments and NPI registry. It is of note that a subset of twenty contributors had multiple states of residency, primary and secondary medical practices located in more than one state, and/or held medical licensure in multiple states. Geographic location was determined by contributors’ state of primary medical practice, and it is thus possible that contributors could represent more than one state each. Future work might also consider examining whether there are racial and ethnic disparities in the composition of contributors, or patient representatives, to the *DSM-6*.

## Conclusion

To ensure the *DSM* is representative of the population in which it serves, a diverse group of contributors constituting equal proportions of male and female contributors should be selected for future editions of the *DSM*. The implementation of gender parity among contributors may procure more robust perspectives on current diagnoses. Additionally, advocating for representation of contributors from all regions of the US will facilitate a more diverse and informed perspective on diagnostic criteria, leading to improved patient care.

## Data Availability

All data produced in the present study are available upon reasonable request to the authors.

## Acknowledgements

We thank Lisa Cosgrove, PhD, who served as the senior author on our previous publication, for her contributions to the foundational work that informed this study.

## Contributors

BP conceptualized the paper. AD and LD developed the first draft of the paper, and all authors reviewed and contributed to all subsequent drafts. AD and LD searched the Open Payments database and NPI registry for authors of the *Diagnostic and Statistical Manual of Mental Disorders*, documented all conflicts of interest and demographic information, and conducted literature searches. LD and BP developed all the figures. All authors contributed to the manuscript revision and read and approved the submitted version. The corresponding author attests that all listed authors meet authorship criteria and that no others meeting the criteria have been omitted.

## Funding

No external funding received.

## Competing Interests

All authors have completed the ICMJE uniform disclosure form at www.icmje.org/coi_disclosure.pdf and declare:

AD reports employment by PerkinElmer, working on assignment at GlaxoSmithKline (2023-Present), outside the submitted work; BP reports contributing to an osteoarthritis research team supported by Pfizer and Eli Lilly (2019-21) and reports receiving grants from the Pennsylvania Academic Clinical Research Center, outside the submitted work; no support from any organization for the submitted work; no financial relationships with any organizations that might have an interest in the submitted work in the previous three years; no other relationships or activities that could appear to have influenced the submitted work.

## Notes

### Funding Statement

This study did not receive any funding.

