## Supplementary figures and images for "Identification of pronounced gender and geographic differences in the Diagnostic and Statistical Manual of Mental Disorders, fifth edition, text revision (*DSM-5-TR*)"

### Supplemental Figure 1

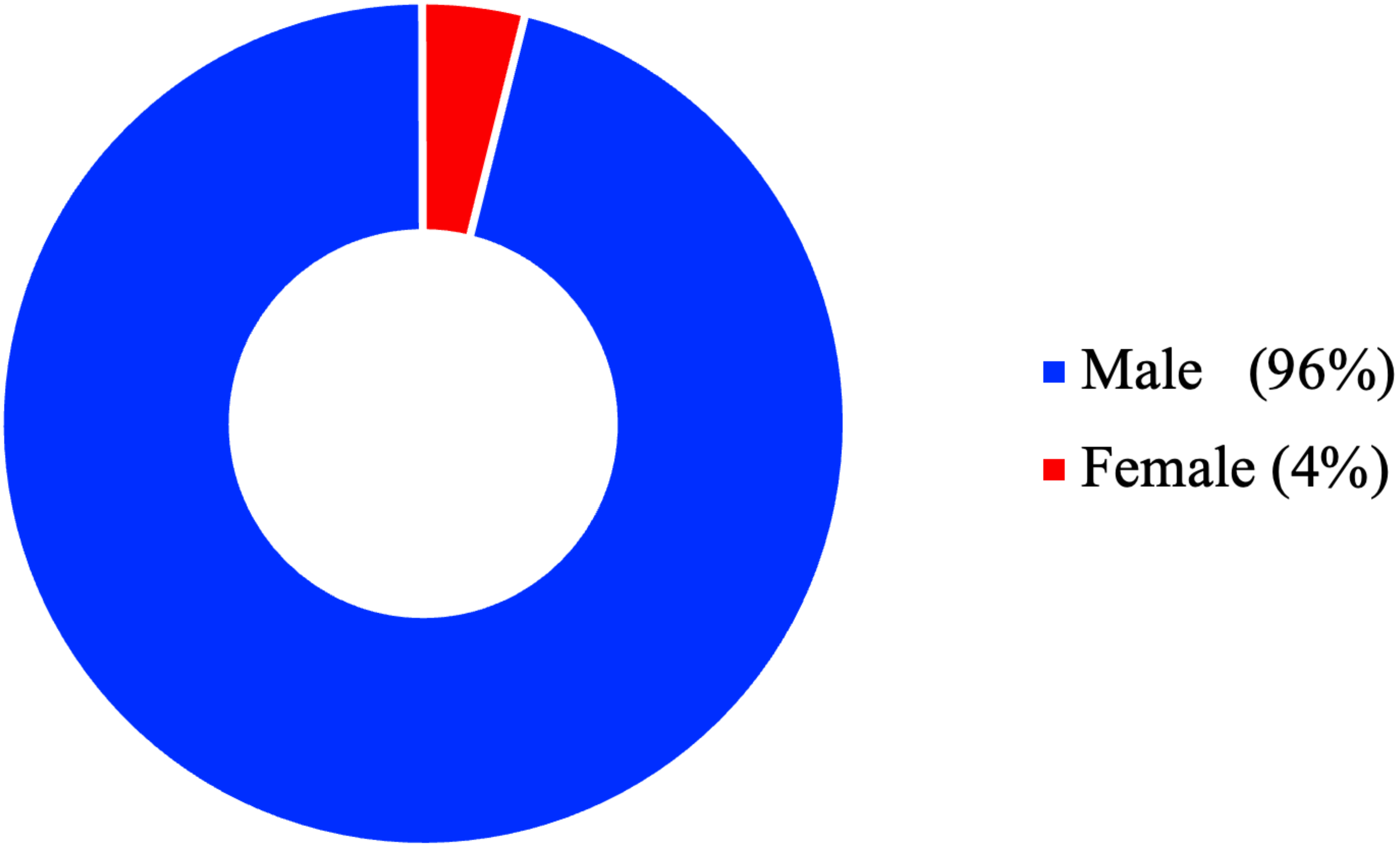
